# Predictive Ability of the Desire to Avoid Pregnancy Scale

**DOI:** 10.1101/2022.10.17.22281028

**Authors:** JA Hall, G Barrett, J Stephenson, CH Rocca, N Edelman

## Abstract

**Study question:** What is the predictive ability of the Desire to Avoid Pregnancy Scale, with regard to pregnancy within one year, and how could it be used as a screening tool?

**Summary answer:** The Desire to Avoid Pregnancy (DAP) Scale is highly predictive of pregnancy within one year and could be used as a screening tool with a suitable cut-point selected according to the purpose.

**What is known already:** There is no existing screening instrument that can reliably predict pregnancy. The Desire to Avoid Pregnancy Scale is a new measure; understanding its sensitivity and specificity as a screening tool for pregnancy as well as its predictive ability and how this varies by socio-demographic factors is important to inform its implementation.

**Study design, size, duration:** A prospective cohort study of 994 non-pregnant women in the UK, with desire to avoid pregnancy measured at baseline and occurrence of pregnancy assessed every quarter for one year. Almost 90% of eligible participants completed follow-up at 12 months; those lost to follow-up were not significantly different on key socio-demographic factors.

**Participants/materials, setting, methods:** The cohort was recruited using social media as well as advertisements in a university, school, abortion clinic and outreach sexual health service. Participants completed an online survey at baseline in October 2018 and every quarter for a year. We used baseline DAP score and a binary variable of whether they had experienced pregnancy during the study to assess the sensitivity, specificity, area under the ROC curve (AUROC) and positive and negative predictive values (PPV and NPV) of the DAP at a range of cut-points. We also examined how the predictive ability of the DAP varied according to socio-demographic factors and by the time frame considered (e.g., pregnancy within 3, 6, 9 and 12 months).

**Main results and the role of chance:** At a cut-point of <2 on the 0-4 range of the DAP scale, scale score had a sensitivity of 0.78 and specificity of 0.81 and an excellent AUROC of 0.87. In this sample the prevalence of pregnancy was 16% (95% confidence interval (CI) 13%, 18%) making the PPV 43% and the NPV 95% at this cut-point. The DAP score was the factor most strongly associated with pregnancy, even after age and number of children were taken in to account, with a 78% reduction in the odds of pregnancy for every one-point increase in the DAP (Odds Ratio 0.22 95% CI 0.17, 0.29). The association between baseline DAP score and pregnancy did not differ across time frames.

**Limitations, reasons for caution:** While broadly in line with the UK population in terms of ethnicity, there were small numbers of pregnancies in participants who were from ethnicities other than white. Further work to explore the DAP in non-white ethnicities and languages other than English that are commonly spoken in the UK, as well as exploring pregnancy preferences by sexuality and in people of all genders, will be important next steps, as we did not ask about gender identity.

**Wider implications of the findings:** This is the first study to assess the DAP scale as a screening tool and shows that its predictive ability is superior to the limited pre-existing pregnancy prediction tools. Based on our findings, the DAP could be used with a cut-point selected according to the purpose.

**Study funding/competing interest(s):** The study was funded by an NIHR Advanced Fellowship held by JH (PDF-2017-10-021). The authors declare that they have no conflicts of interest.

**Trial registration number:** n/a

## Introduction

A longstanding gap in the reproductive health field has been the availability of a screening instrument that can reliably predict a person’s likelihood of becoming pregnant. Such an instrument would be of particular use for researchers either trying to identify a specific cohort (e.g. for preconception research) or conducting research for which it is important to exclude participants who are likely to become pregnant, (e.g. some pharmacological studies). The Desire to Avoid Pregnancy (DAP) scale is a psychometric instrument that measures a person’s preferences about a potential future pregnancy. The DAP was developed as a research instrument in the USA^1^ and was validated for use in the UK in 2022^2^. This 14-item tool was shown to have good predictive ability for pregnancy occurring within one year in the UK. Women^a^ with the lowest desire to avoid pregnancy (a score of zero) had an 80% chance of becoming pregnant within 12 months compared to <1% in women with the highest desire to avoid pregnancy (a score of four). To our knowledge the DAP scale is the only purposively designed and evaluated prospective measure of pregnancy preferences.

The DAP is a new measure and understanding its potential use as a screening tool and ability to predict pregnancy is important for both research and clinical purposes. While the overall predictive ability over 12 months appears to be high, further exploration is required to understand its sensitivity and specificity relative to incident pregnancy, and whether the predictive ability of the DAP varies according to sociodemographic factors, as well as over different timeframes^3-5^.

The aims of this paper are to: 1) to examine the sensitivity and specificity of the DAP scale relative to incident pregnancy; and 2) to explore the scale’s predictive ability across socio-demographic characteristics and time frames.

## Materials and methods

### Sample

This analysis was conducted on a cohort of 994 non-pregnant participants recruited in October 2018 and followed up for one year. The full details of recruitment and participation are described elsewhere^2^ but, in brief, people who self-reported as female, were pre-menopausal and not sterilised, aged 15 or over and living in the United Kingdom, were recruited though a mixture of site-based advertising (school, university, sexual health and pregnancy termination clinics) and online recruitment through both paid advertisements (Instagram and Facebook) and sharing through researchers’ and participants’ networks. Participants completed an online RedCap survey at baseline and every three months for 12 months^6,7^ that included the DAP scale and other questions about pregnancy preferences, contraceptive use, preconception preparation and socio-demographics. At each of the three-month follow-up surveys, participants were asked whether they were currently pregnant or had been pregnant since the last survey.

### Measures

#### Outcome

Our outcome was experience of an incident pregnancy over 12 months (yes/no), created using self-reported pregnancy data across all follow-up surveys. For analyses of time frame, we also looked at incident pregnancy by 3, 6, and 9 months, individually.

#### DAP Scale

The DAP scale is a psychometrically validated measure of a woman’s preferences about a potential future pregnancy, developed using an extensive item development process and item response theory to create the final tool^1^. Its 14 items cover three conceptual domains: 1) cognitive desires and preferences; 2) affective feelings and attitudes; and 3) anticipated practical consequences. Each of the 14 items asks respondents to report using a Likert scale on how much they agree or disagree with a statement about either becoming pregnant in the next three months or having a baby in the next year. Each item is scored zero to four; responses are summed and averaged to get a total score between zero and four, with four representing the greatest desire to avoid pregnancy and zero the most open to pregnancy.

### Analysis

#### Sensitivity and specificity of the DAP relative to Pregnancy

The relative importance of sensitivity and specificity vary according to the purpose of using the DAP scale (i.e. whether identifying who will become pregnant (sensitivity) is more important that identifying who will not (specificity) or vice versa), therefore a range of cut-points was explored. Initially the Youden index was used to suggest an empirically optimal cut-point, i.e. the best balance of sensitivity and specificity^8^. This cut point was used to classify women as ‘test positive’ if their score was below the cut-point and was compared with the ‘true positive’ of whether they experienced a pregnancy between baseline and 12months. The sensitivity, specificity, area under the receiver operator curve (AUROC) and positive and negative predictive values (PPV and NPV) were then calculated. The AUROC represents the DAP’s ability to discriminate between those who will and will not become pregnant, where 0.5 is no better than random and 1.0 is perfect discrimination. An AUROC of 0.7-0.8 was considered acceptable, 0.8-0.9 excellent and >0.9 outstanding^9^. This process was then repeated for a range of cut-points to provide information to enable the selection of the most suitable cut-point depending on purpose.

#### Predictive ability of the DAP

##### Univariate analysis

Univariate analyses were conducted to explore how pregnancy preferences, as measured by DAP score, vary by age, ethnicity, education, number of children and relationship status, using the Kruskall-Wallis test for ordered categorical variables (where DAP score was not expected to increase or decrease consistently (age group, number of children in the household, ethnicity)) and the Kendall’s tau where it was (education, relationship status). Non-parametric tests were used given the non-normal distribution of the DAP score. Baseline data were used for all socio-demographic factors as there was minimal change over follow up. The relationship between each factor and occurrence of pregnancy was also examined with logistic regression. We examined differential attrition by socio-demographics and baseline DAP score using t-tests, Kruskall-Wallis and chi-squared tests, as appropriate.

##### Multivariable analysis

A multivariable logistic regression model of DAP score as a predictor of pregnancy was created by including all the socio-demographic factors considered and removing them in a manual backwards stepwise process, starting with the largest p-value and retaining only variables where p<0.1. The predicted probabilities of pregnancy from this model were examined, both overall and by age and number of children.

##### Timeframes

To determine whether the predictive ability of DAP score varies by the time frame considered, and therefore confirm whether asking preferences annually is sufficient or should be done more frequently, the baseline DAP score was used to calculate the odds of pregnancy between baseline and three, six and nine months respectively using logistic regression. Given low attrition and to ease interpretation, we include participants in pregnancy denominators until they were lost from the cohort and report percentages rather than rates.

### Ethical approval

Ethical approval was granted by the UCL Research Ethics Committee (ref 3974.003).

## Results

### Samples

As described previously^2^, the baseline cohort of 994 women were aged 15-50 years (median 31, IQR 23-36, mean 29.7). Most were white (84%), described themselves as heterosexual (82%) and were in a relationship (82%). Over half (57%) had one or more children in the household, 25% had completed secondary school, 39% had an undergraduate degree and 31% had postgraduate or other professional qualifications. Almost 90% (831/929) of participants eligible to take part in the 12month survey did so. The women who did not take part in the 12month follow up survey did not differ by age, ethnicity, relationship status, number of children or baseline DAP score.

### Sensitivity, specificity of the DAP

Over the 12month study 14.0% (139/994) of women experienced pregnancy. The Youden recommended cut-point was 1.96 (rounded here to <2), at which point the DAP had a sensitivity of 0.78 and specificity of 0.81. The AUROC was excellent (0.87) (Figure 1), suggesting that DAP score is a good discriminator of whether someone will become pregnant in the next 12months. The sensitivity tells us that 78% of all the women who will become pregnant over the next 12months would be detected by using the DAP with a cut-point of <2. In this sample the prevalence of pregnancy was 16% (95%CI 13%, 18.3%) making the PPV 43% and the NPV 95% at this cut-point. The PPV shows us that, in this sample, 43% of people with score of <2 will become pregnant within 12 months.

**Figure 1.**
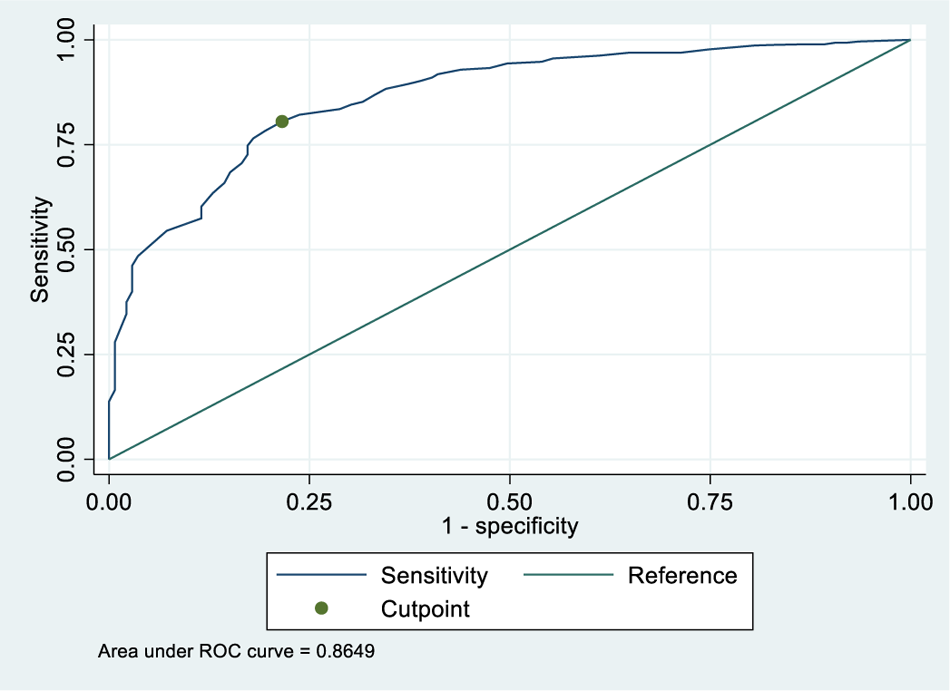
Receiver operating characteristic (ROC) curve for DAP score at cut-point <2

Depending on the population and the purpose of the question the cut-point could be adapted to suit the purpose, as shown in Table 1. For example, if the identification of a preconception cohort was the goal a lower cut-point, such as 0.5, yields a PPV of 73%, i.e., 73% of people with score of <0.5 will become pregnant within 12 months. Conversely, using a cut-point of 3, <1% of people scoring over 3 will become pregnant within 12 months.

**Table 1.** Sensitivity, specificity, AuROC, positive and negative predictive values at a range of DAP cut-points

| Cut-point | <0.5 |  |  | <1 |  |  | <2 |  |  | <3 |  |  |
| --- | --- | --- | --- | --- | --- | --- | --- | --- | --- | --- | --- | --- |
|  | Estimate | 95%CI |  | Estimate | 95%CI |  | Estimate | 95%CI |  | Estimate | 95%CI |  |
| Sensitivity | 17.3% | 11.4% | 24.6% | 50.4% | 41.8% | 58.9% | 78.4% | 70.6% | 84.9% | 97.1% | 92.8% | 99.2% |
| Specificity | 98.8% | 97.7% | 99.4% | 94.4% | 92.5% | 95.9% | 80.5% | 77.5% | 83.3% | 46.2% | 42.5% | 49.8% |
| AuROC | 0.58 | 0.55 | 0.61 | 0.72 | 0.68 | 0.77 | 0.80 | 0.76 | 0.83 | 0.72 | 0.69 | 0.74 |
| Positive predictive value | 72.7% | 54.5% | 86.7% | 62.5% | 52.9% | 71.5% | 42.9% | 36.7% | 49.2% | 25.2% | 21.6% | 29.1% |
| Negative predictive value | 86.5% | 84.0% | 88.7% | 91.1% | 88.8% | 93.0% | 95.2% | 93.3% | 96.8% | 98.9% | 97.1% | 99.7% |

### Predictive ability of the DAP

#### Univariate analysis

As previously reported^2^, univariate analysis showed that for every one-point increase in DAP score the odds of pregnancy within 12 months decreased by 78% (OR 0.22, 95%CI 0.17, 0.28). Women with a DAP score of zero had a predicted 79.4% chance of pregnancy in the next 12 months, while 0.89% of those with a DAP score of four experienced pregnancy.

Preferences regarding future pregnancy varied by all five socio-demographic factors, as shown in Table 2. Desire to avoid pregnancy was highest in; 15-19 age group; those not in a relationship; those with three or more children; women in Black, Asian, Mixed and Other ethnic groups; women whose highest completed level of education was secondary (high) school (usually school up to age 18).

**Table 2.** Distribution of DAP score by socio-demographic variables

|  | DAP Score |  |  |  |  |
| --- | --- | --- | --- | --- | --- |
|  | N | Median | Interquartile range |  | p value |
| Age group (years) |  |  |  |  |  |
| 15-19 | 139 | 3.50 | 3.00 | 3.86 | <0.001 |
| 20-24 | 143 | 3.14 | 2.36 | 3.64 |  |
| 25-29 | 139 | 2.29 | 1.57 | 2.93 |  |
| 30-34 | 224 | 2.14 | 1.11 | 2.86 |  |
| 35-39 | 209 | 2.57 | 1.57 | 3.21 |  |
| >=40 | 101 | 2.79 | 2.14 | 3.36 |  |
| Missing | 39 | 2.71 | 2.14 | 3.29 |  |
| Relationship status |  |  |  |  |  |
| Married / civil partnership | 479 | 2.36 | 1.36 | 3.00 | <0.001 |
| In a relationship, not married | 338 | 2.82 | 2.07 | 3.57 |  |
| Not in a relationship | 152 | 3.36 | 2.86 | 3.79 |  |
| Missing | 25 | 2.86 | 2.29 | 3.29 |  |
| Number of children in the household |  |  |  |  |  |
| 0 | 430 | 2.07 | 3.00 | 3.71 | <0.001 |
| 1 | 208 | 0.86 | 1.89 | 2.61 |  |
| 2 | 233 | 2.07 | 2.79 | 3.29 |  |
| >=3 | 78 | 2.29 | 2.86 | 3.43 |  |
| Missing | 45 | 2.29 | 2.93 | 3.29 |  |
| **Grouped ethnicity** |  |  |  |  | 0.042 |
| White | 834 | 1.79 | 2.64 | 3.36 |  |
| Black, Asian, Mixed and Other ethnicities | 128 | 2.11 | 2.82 | 3.64 |  |
| Missing | 32 | 2.29 | 2.89 | 3.50 |  |
| **Highest completed level of education** |  |  |  |  | <0.001 |
| School* | 247 | 2.57 | 3.29 | 3.79 |  |
| Undergraduate | 389 | 1.57 | 2.50 | 3.14 |  |
| Postgraduate or professional qualifications | 309 | 1.71 | 2.50 | 3.14 |  |
| Missing | 31 | 2.14 | 2.93 | 3.64 |  |

\* Includes those currently still at school (n=17) and one person with no qualifications

Likelihood of pregnancy in the next 12 months also varied by baseline measures of age, relationship status, number of children in the household and education level, but not by ethnicity. Pregnancy in the next 12 months was most likely to occur in the 30-34 age group compared to women aged 15-19 (OR 12.9 95%CI 3.9, 42.2), in women in a marriage or civil partnership compared to those who were not in a relationship (OR 9.55 95%CI 3.44, 26.5), women with one child in the household compared to women with none (OR 5.07 95%CI 3.22, 7.97) and women with completed undergraduate level educational attainment compared to those whose highest level of education was school (OR 4.61 95%CI 2.44, 8.71).

#### Multivariable analysis

In the development of the multivariable model, relationship status (p=0.50), ethnicity (p=0.43) and education level (p=0.20) were not significantly associated with pregnancy when all factors were included. Only age and number of children remained in the final multivariable model. The relationship between DAP score and pregnancy was unchanged in the multivariable model and was the strongest predictor, as shown in Table 3.

**Table 3.**
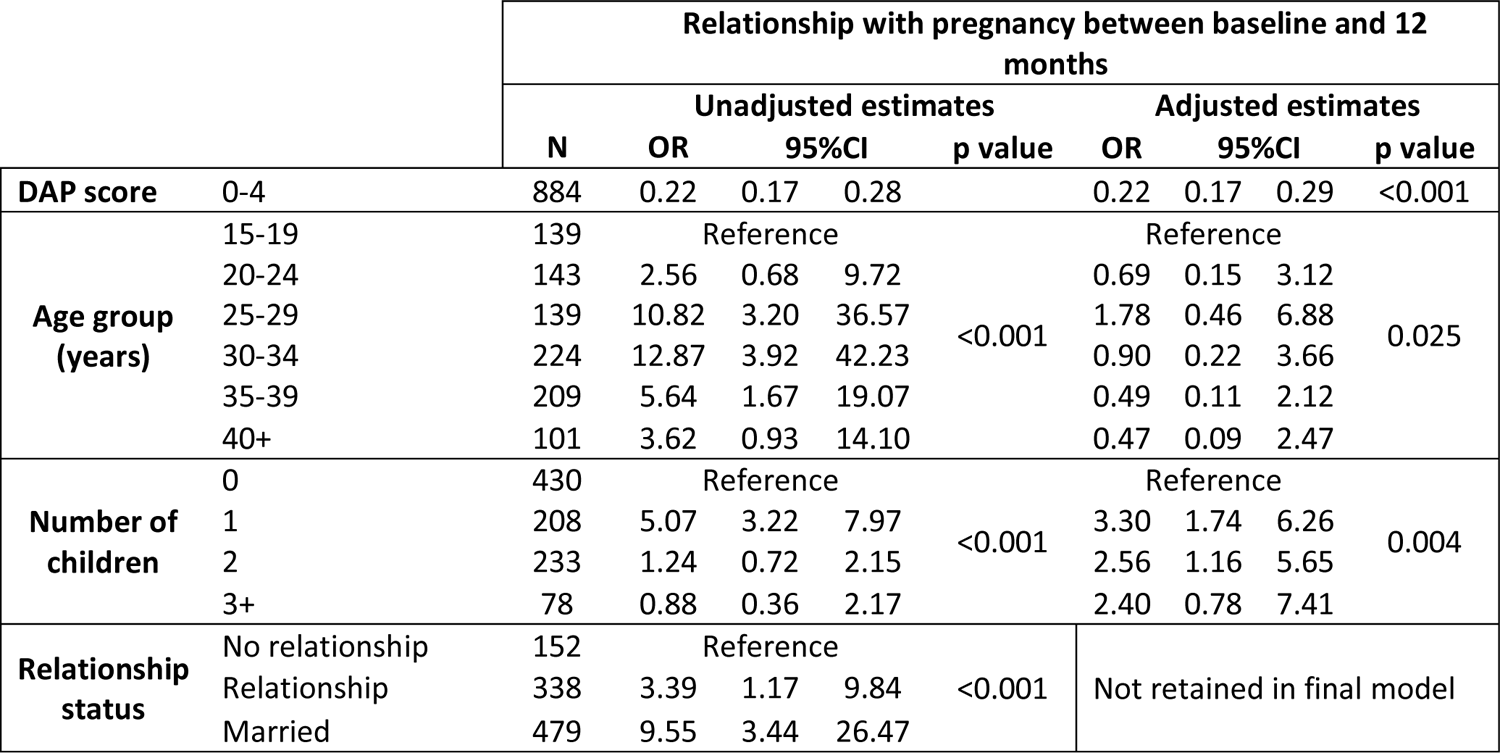
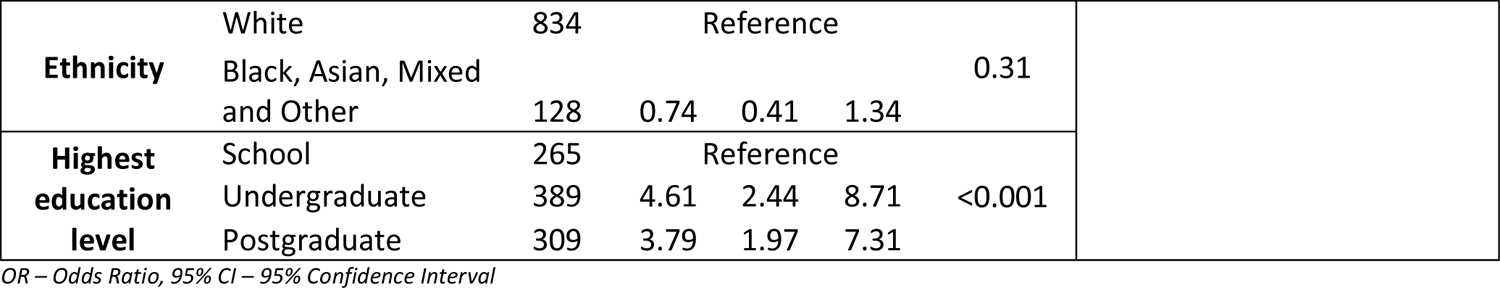
Multivariable logistic regression of the odds of pregnancy within 12 months.

The predicted probabilities of pregnancy according to selected age groups and number of children estimated by the multivariate model can be seen in Figure 2. The probability of pregnancy was highest (87.4%) in women aged 25-34 who already had one child in the household and scored zero on the DAP score at baseline (indicating that they desired a pregnancy). Regardless of age or number of children, women with a DAP score of four at baseline were very unlikely (<2%) to have a pregnancy within the next year. Women who were aged 35 and over with no children but who scored zero on the DAP at baseline had a 54.4% chance of pregnancy.

**Figure 2.**
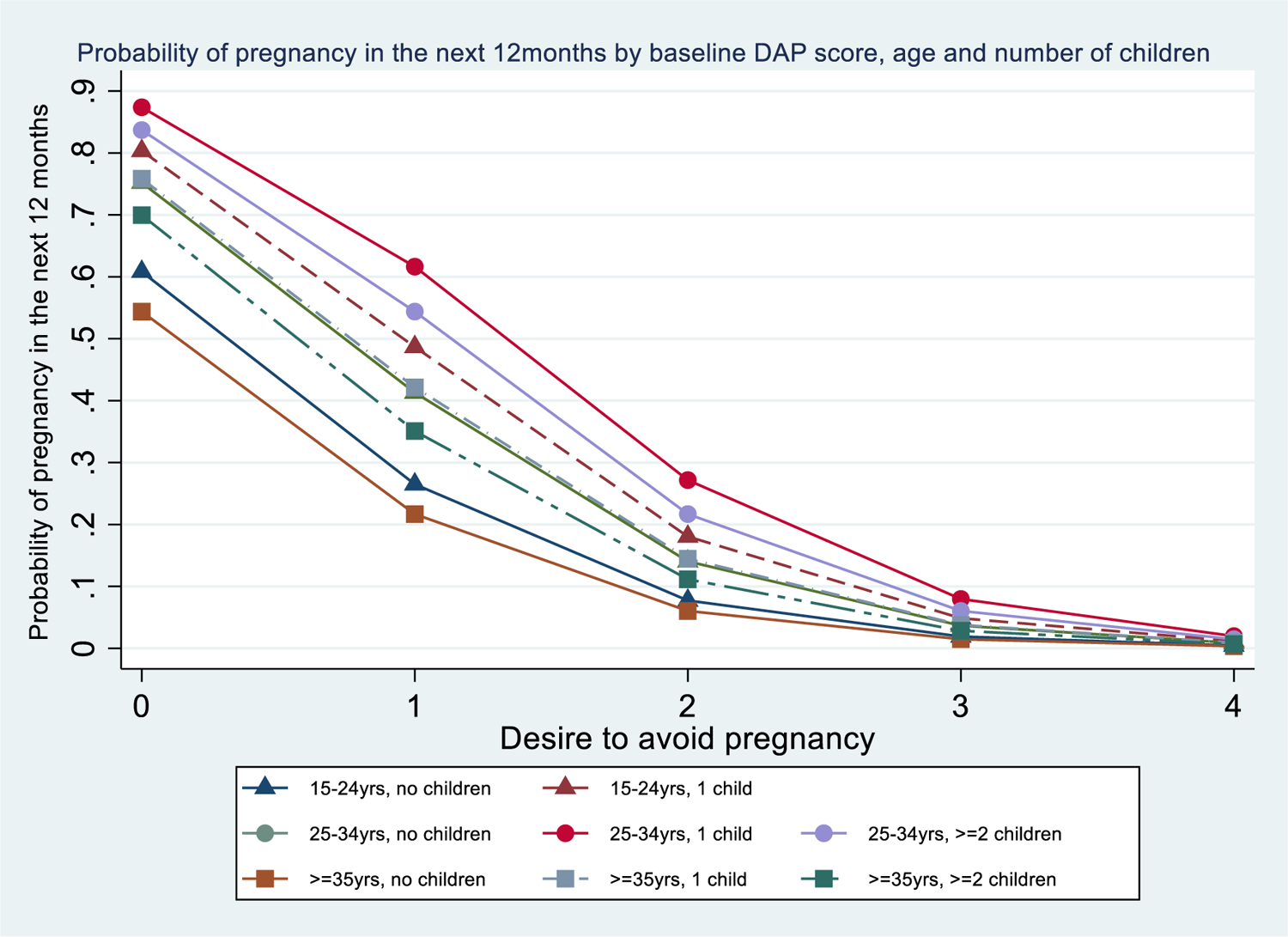
Predicted probability of pregnancy within 12 months based on DAP score taking into account age group and number of children in the household.

#### Time frames

The association between baseline DAP score and pregnancy did not differ across time points as shown in Table 4.

**Table 4.** Odds of pregnancy (OR) at each follow-up by baseline DAP score with 95% confidence interval (95%CI)

| Relationship between baseline DAP score and pregnancy at: |  |  |  |
| --- | --- | --- | --- |
|  | OR | 95%CI |  |
| 3 months | 0.24 | 0.17 | 0.34 |
| 6 months | 0.19 | 0.14 | 0.26 |
| 9 months | 0.18 | 0.13 | 0.24 |
| 12 months | 0.22 | 0.17 | 0.28 |

## Discussion

This is the first study to examine the potential utility of the DAP scale as a tool to predict future pregnancy. Based on this analysis, the DAP could be used, with a cut-point selected based on the purpose, to identify who is likely to become pregnant over the next 12 months and who is not.

While pregnancy desire and occurrence are associated with a range of socio-demographic factors, both in our data and the wider literature^3-5^, the DAP score is the most strongly associated factor, based on the size of the odds ratio, even when other factors are taken into account. There were differences in the probability of pregnancy in those with a DAP score of zero based on age and number of children, in keeping with the literature^3,4,10^ and almost no difference in the probability of pregnancy, regardless of age or number of children, in those with the highest desire to avoid pregnancy. This demonstrates the gap between wanting something and the ability to make it happen which, in the case of pregnancy, is only within a person’s control to some extent. Pregnancy may be further affected by external factors such as fecundability and the supportability of pregnancy, defined by MacLeod as ‘the capacity of a woman to carry a pregnancy in such a way that she experiences positive health and welfare’^11^. However, the overall predicted probability of pregnancy was remarkably congruent with evidence that approximately 80% of couples who desire pregnancy will conceive within 12 months^12,13^.

There have been few attempts to develop predictive models for pregnancy, and many of those have focused on sub-fertile couples^14-16^. AUROCs for pregnancy prediction models in reproductive health have generally been low, ranging from 0.56-0.67, which may in part reflect the challenges of predicting pregnancy in a heterogeneous sub-fertile population^17^. One study, of women who were trying to conceive and who enrolled in a preconception health study, used machine learning to develop a prediction model from a pool of 163 potential predictors encompassing socio-demographics, diet and lifestyle, medical history and some partner characteristics^10^. The authors developed multiple models, considering different time frames and populations, with AUROCs between 0.65-0.71. In comparison to these other models, the 14-item DAP’s AUROC of 0.87 is very high. This is especially noteworthy as participants were not all trying to conceive and therefore may be more representative of the general population and of how the DAP may perform in practice.

The cut-point, and therefore sensitivity and specificity, could be varied by setting and purpose for asking the DAP. For example, in the context of a screening tool in primary care to identify who would benefit from preconception advice a lower cut-point could be used as it would be important not to have too many false positives (i.e., women predicted to get pregnant who will not) as this could overload services and be unsustainable and unacceptable. Alternatively, for researchers planning a trial of a teratogenic agent and wishing to reduce attrition, one might select a high cut-point to ensure the lowest proportion of participants experience pregnancy.

## Strengths and limitations

The analysis has been conducted on a large cohort with little loss to follow-up (the women lost were not significantly different to those retained, suggesting that there is no selection bias in the loss to follow up). While a non-probability sample, comparison of the socio-demographics of the cohort indicate that it is broadly representative of the UK population^2^, suggesting generalisability of our findings to a primary care population. However, the cohort was slightly more educated than average, which is common with online surveys, indicating potential selection bias. Importantly the cohort was neither sub-fertile nor self-identified as preconception, a distinction from previous pregnancy prediction model cohorts. We have demonstrated the high discrimination of the full DAP scale in excess of previously developed pregnancy prediction models.

While we included a range of sociodemographic factors known to be associated with pregnancy preferences in our multivariable model, we did not include all factors associated with fertility, such as BMI or smoking^18^ as we did not have data on these. Inclusion of these variables may have strengthened the model. Assessing the DAP scale’s performance in other languages, cultural settings and exploring pregnancy preferences by sexuality and in people of all genders will be important next steps.

The DAP scale could be useful clinically in a range of self-completion or digital formats to identify who is likely to become pregnant and who is not. Further work should be done to develop a clinical tool that maximises discrimination while being practical within a face-to-face clinical encounter.

## Conclusion

This study shows the excellent predictive ability of the DAP, which was the strongest predictor of pregnancy even when other socio-demographic factors were taken into account. The estimates of the predicted probability of pregnancy using the DAP score are stable, suggesting it may not need to be asked more than once a year. At a cut-point of <2 sensitivity and specificity are optimised at 78% and 81% respectively, however the information on the sensitivity and specificity of the DAP at a range of cut-points will support academics and clinicians to adapt the choice of cut-point according to their aim and needs.

## Data Availability

The dataset is available in the UCL Research Data Repository.

## Author’s contributions

JH: guarantor, conceptualisation, funding acquisition, methodology, investigation, data curation, formal analysis, writing-original draft, visualisation, writing-review and editing, project administration. GB: conceptualisation, methodology, writing-review and editing. JS: conceptualisation, writing-review and editing, supervision CR: conceptualisation, methodology, writing-review and editing. NE: interpretation, writing-review and editing

## Footnotes

a Where we refer to ‘women’ this should be taken to include people who do not identify as women but who have the capability to become pregnant.

